# Nutritional Status and Associated Factors Among Children Aged 6-24 Months at a Primary Health Care Centre in Conflict-Affected Gaza

**DOI:** 10.64898/2026.05.12.26353044

**Authors:** Lina Murtaja, Hamza Abdeljawad, Ahmed Najim, Josie Rodgers, Reham Almukbel, Kinan Mokbel

## Abstract

**Background/Objectives:** Children aged 6-24 months are highly vulnerable to malnutrition during conflict because they depend on breastfeeding, complementary feeding and functioning nutrition services. This study assessed nutritional status, socioeconomic correlates, maternal knowledge and primary health care centre (PHCC) nutrition service gaps in Gaza.

**Subjects/Methods:** This cross-sectional study was conducted at Al-Daraj Martyrs Health Centre, one of the remaining functioning PHCCs in Gaza City during the study period, between late August and October 2025. Mother-child pairs were recruited by convenience sampling. Of 276 approached, 200 were included after non-response and exclusion of questionnaires with missing anthropometric data. Data came from structured interviews and medical records; haemoglobin results were available for 55 children.

**Results:** Stunting affected 12.5% of children, underweight 20.1%, wasting 20.8%, and anaemia 63.6% of the haemoglobin-tested subsample. Underweight was associated with household food shortage (*p*=0.013) and previous malnutrition treatment (*p*=0.002), wasting with child age category (*p*=0.0024), and anaemia with paternal unemployment (*p*=0.020). Maternal knowledge and practice scores were positively correlated (r=0.177, *p*=0.012), but neither was independently associated with stunting or underweight in adjusted models. PHCC nutrition support was limited, with 71.0% of mothers reporting nurse-provided nutrition advice and 52.5% reporting growth-chart review.

**Conclusions:** In this clinic-based sample from conflict-affected Gaza, malnutrition among children aged 6-24 months was substantial. The overall pattern suggests that nutritional risk was shaped more by structural deprivation and weakened PHCC support than by maternal knowledge alone. These findings underline the need to restore essential nutrition services and improve access to adequate food for young children.

## Introduction

Child malnutrition remains a critical global public health challenge, and recent multi-country analyses from low- and middle-income settings continue to show a substantial burden of stunting, wasting and underweight among children under five (1). These forms of undernutrition remain strongly patterned by socioeconomic conditions and continue to represent a major cause of preventable morbidity and developmental loss in early life. The period from 6 to 24 months represents a critical window of vulnerability, coinciding with complementary feeding introduction and rapid neurodevelopmental maturation (2,3). Nutritional deficits during this period carry lifelong consequences for physical growth, cognitive development, and disease susceptibility (4,5).

The Gaza Strip exemplifies extreme humanitarian crisis conditions with profound impacts on child nutrition. Since October 2023, the escalation of hostilities has created catastrophic deterioration in food security and healthcare access (6,7). Recent assessments document severe food insecurity and sharp reductions in access to meat, dairy, fruit, and vegetables in Gaza (8). Such dietary collapse implies not only reduced energy intake, but also profound deterioration in dietary adequacy, with loss of access to protein-rich and micronutrient-dense foods essential for infant growth and development. For children aged 6-24 months, who depend on nutrient-dense complementary feeding alongside breastfeeding, this pattern is likely to increase the risk of deficiencies affecting linear growth, immune function, and neurodevelopment (3,5).

Pre-conflict Gaza already harboured substantial nutritional vulnerabilities, including 35.6% anaemia prevalence among children aged 24-59 months and evidence of suboptimal breastfeeding knowledge and practices among mothers (9,10). Importantly, pre-conflict household assessments in Gaza also documented food insecurity, dietary inadequacy, and malnutrition among refugee children and their households, suggesting that the current emergency has intensified a pre-existing nutritional fragility rather than creating it de novo (11). This was consistent with broader Eastern Mediterranean evidence showing persistent concerns around child dietary adequacy and nutritional status across the region (12). The current crisis has likely intensified these baseline challenges substantially. Recent surveillance in Gaza reported that children aged 6 to <24 months had the highest prevalence of acute malnutrition in an under-5 sample, with 11.0% classified as severe acute malnutrition and 17.9% as moderate acute malnutrition on Mid-Upper Arm Circumference (MUAC)-based screening (13).

Healthcare infrastructure has sustained severe damage, with primary health care centres (PHCCs) operating under extreme constraints (14). Despite the recognised role of growth monitoring and caregiver feeding support in child-health programmes (15), capacity has been critically compromised by staff shortages, interrupted supply chains, and equipment destruction (14,16). Micronutrient supplementation coverage has also been severely disrupted, including routine vitamin A supplementation and other preventive nutrition services (16), while recent reports of severe dietary deterioration in Gaza suggest worsening maternal and infant micronutrient adequacy (8). Recent expert commentary has also warned that infant formula-centred responses in Gaza may be harmful if untargeted distributions displace breastfeeding in settings with unsafe water and inadequate hygienic preparation conditions (17). Recent humanitarian reporting also indicates that conflict escalation, displacement orders, and access restrictions have disrupted preventive nutrition services and worsened food insecurity and malnutrition conditions in parts of Gaza (18,19). Deficiencies in these micronutrients are associated with impaired foetal and infant growth, reduced immune resilience, and increased risk of anaemia and undernutrition (3,5).

Maternal knowledge and feeding practices are recognised determinants of child nutrition in more stable settings (20,21). In humanitarian crises, however, their protective effect may be limited when food access, displacement, insecurity, and disrupted services constrain caregiver choice (8,14,16). Limited evidence examines these relationships in acute conflict settings, particularly during severe conflict-related food insecurity or catastrophic humanitarian conditions. This creates an important operational and evidence gap. Existing Gaza data describe either pre-conflict nutritional vulnerability, broad household food insecurity, or acute malnutrition across wider paediatric age ranges, but do not adequately characterise the 6-24-month group within functioning PHCC settings during active conflict (8,9,10,13,14,16). That gap matters because nutritional deterioration during the complementary-feeding period can progress rapidly and because preventive opportunities at primary care level are highly time-sensitive in this age group (2,3,5,22,23).

Direct evidence on malnutrition among children aged 6-24 months in Gaza during the current crisis remains limited (13). The 6-24-month cohort, representing peak malnutrition vulnerability and intervention responsiveness, requires urgent focused investigation (22,23). This matters because children aged 6-24 months are both biologically vulnerable and highly dependent on timely preventive contact with primary care, making PHCC-based evidence especially important when population-level surveillance becomes difficult during conflict (2,3,16). This was especially relevant during the study period, when Al-Daraj Martyrs Health Centre was one of the remaining functioning PHCCs in Gaza City, making evidence from this site particularly important for understanding child nutrition and residual PHCC service capacity under active conflict conditions. Accordingly, this study was designed not only to describe malnutrition prevalence in children aged 6-24 months during active conflict, but also to generate clinically and operationally relevant evidence for humanitarian response. Specifically, it examined socioeconomic and demographic factors associated with malnutrition outcomes, assessed PHCC nutrition service delivery, and evaluated whether maternal knowledge and reported practices remained associated with child nutritional status under conditions of extreme structural deprivation. To our knowledge, this is among the first clinic-based studies to integrate anthropometric outcomes, haemoglobin-based anaemia, maternal knowledge and practice, PHCC nutrition service delivery and adjusted modelling of undernutrition risk among children aged 6-24 months in conflict-affected Gaza.

## Materials And Methods

### Study design and setting

This cross-sectional study was conducted at Al-Daraj Martyrs Health Centre in Gaza City between late August and October 2025. During the study period, the centre was one of the remaining functioning PHCCs in Gaza City, giving the site particular operational importance as a remaining access point for routine child health services.

### Participants, eligibility, and sampling

The target population comprised children aged 6 to 24 months attending the centre with their mothers. Children with congenital anomalies, extreme prematurity (<28 weeks’ gestational age), or severe chronic illnesses judged at recruitment likely to substantially affect nutritional status were excluded. A small number of children nevertheless had chronic conditions recorded in the routine clinical data and were retained as a descriptive covariate because these had not been treated as exclusionary at recruitment.

Based on clinic attendance records from August 2025, 977 children aged 6 to 24 months attended the centre and formed the sampling frame. Sample size was estimated using Epi Info StatCalc, which yielded a target of 276 mother-child pairs. A convenience sample was then recruited. In total, 276 mother-child pairs were approached, of whom 26 were non-respondents, leaving 250 completed questionnaires. A further 50 questionnaires were excluded because both height and weight data were missing. The final analytic sample therefore comprised 200 mother-child pairs. Participant flow through recruitment, exclusions, and the final analytic sample is summarised in Figure 1.

**Figure 1.**
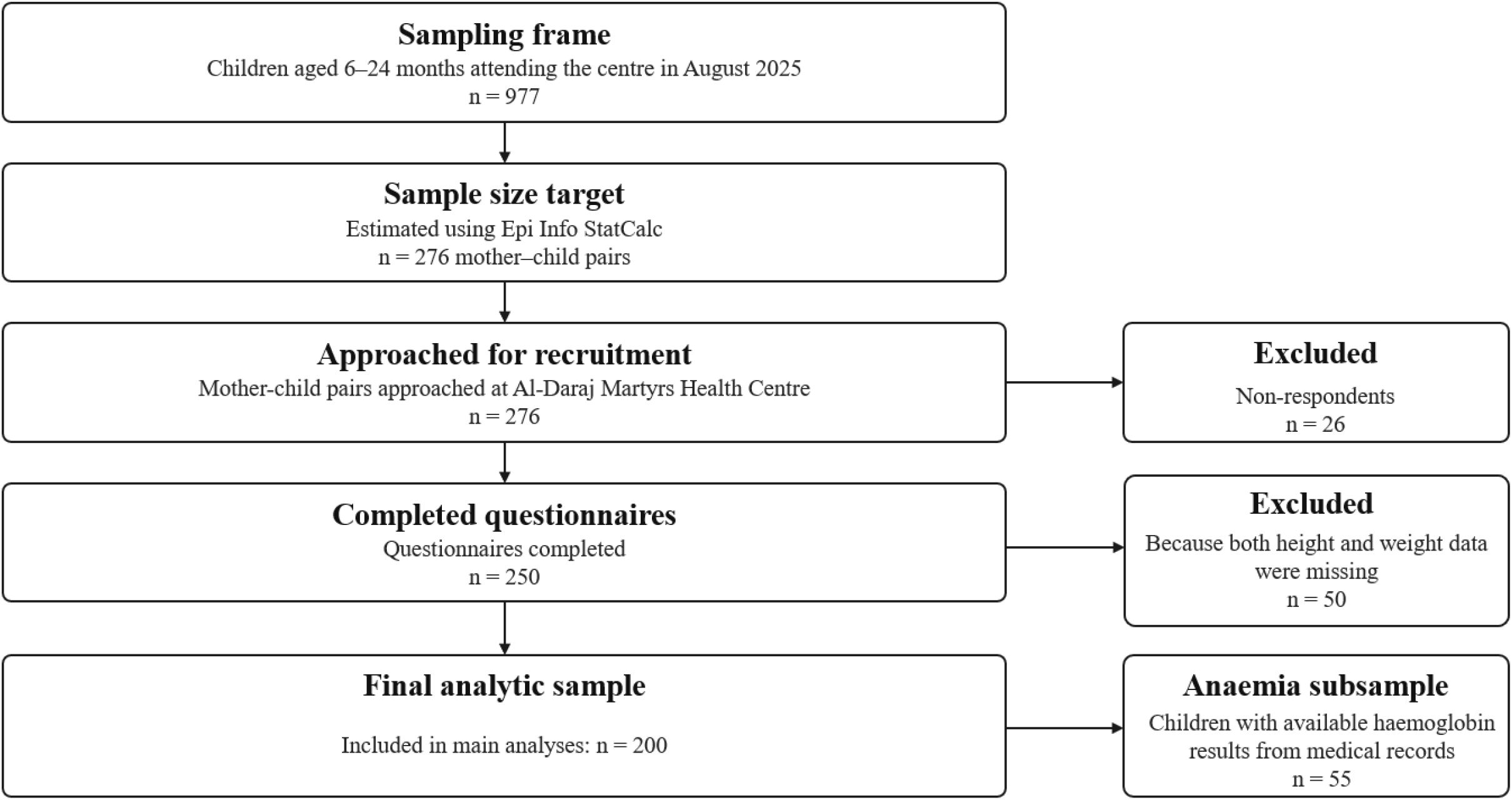
Participant flow diagram Participant flow diagram of study recruitment, exclusions and the final analytic sample.

### Data collection and study measures

Data were collected through structured face-to-face interviews conducted by the principal investigator, with each interview lasting approximately 15 to 20 minutes. The questionnaire covered child feeding, maternal knowledge and feeding practices, household socioeconomic circumstances, and nutrition-related health-service exposure. It also recorded chronic disease and previous malnutrition treatment. The full questionnaire in English and Arabic is provided in the Supplementary Information.

Household food shortage was measured as a binary self-report variable using the question, “Does the family face food shortages?” with response options yes or no. Regular PHCC attendance was also assessed by maternal self-report using the item, “Do you visit the child health center regularly?” with response options yes or no.

### Questionnaire development, reliability, and scoring

Questionnaire development involved a literature review and expert panel evaluation to assess content validity. Pilot testing with 25 mothers was undertaken to assess clarity and feasibility. Internal consistency reliability was acceptable, with Cronbach’s alpha values of 0.740 for the knowledge domain, 0.780 for the practice domain, and 0.776 overall. Detailed reliability results are presented in Supplementary Table S1. These reliability findings supported use of the retained knowledge and practice scores in the main analyses.

The questionnaire initially comprised 14 knowledge items and 13 practice items. Internal consistency of the knowledge and practice scales was assessed using Cronbach’s alpha and item-level diagnostics, including alpha if item deleted. Based on these analyses, three knowledge items and one practice item were removed before calculating the final composite scores. The final knowledge score was based on 11 items, with correct responses scored as 1 and incorrect responses scored as 0, giving a total possible range of 0 to 11. Three negatively phrased knowledge items were reverse-coded. The final practice score was based on 12 items rated on a 5-point Likert scale, with one negatively phrased item reverse-coded, giving a total possible range of 12 to 60. The retained knowledge score corresponded to questionnaire items on colostrum, exclusive breastfeeding, signs of milk adequacy, breastfeeding during infant illness, positioning and latch, breastfeeding alongside food until 24 months or more, complementary feeding at 6 months, food consistency progression, avoiding sugar/salt, food hygiene, and avoiding sugary or energy drinks. The retained practice score excluded the colostrum practice item.

### Anthropometric and haemoglobin data

Anthropometric data, including weight and length or height, were extracted from children’s medical records. These measurements had been recorded routinely by PHCC staff as part of clinical care, although independent verification of equipment calibration and adherence to WHO procedures was not possible. Height-for-age, weight-for-age, and weight-for-height Z scores were calculated using WHO Anthro version 3.2.2 (WHO, Geneva, Switzerland). Nutritional status was classified according to the WHO Child Growth Standards (24), with stunting defined as height-for-age Z score below −2 SD, wasting as weight-for-height Z score below −2 SD, and underweight as weight-for-age Z score below −2 SD.

Haemoglobin levels were obtained from available laboratory test results recorded in the medical files. These data were available for 55 children, representing 27.5% of the analytic sample. Because haemoglobin testing was not available for all participants, the anaemia analysis was based on a selective subsample and should be interpreted cautiously. Anaemia was defined as haemoglobin concentration below 11 g/dL according to WHO criteria (25).

### Statistical analysis

Data were analysed using Stata/SE version 19.0 (StataCorp, College Station, TX, USA). Descriptive statistics were presented as frequencies and percentages for categorical variables and means with standard deviations for continuous variables. Child age was analysed in five categories: 6 months, 7 to 9 months, 10 to 12 months, 13 to 18 months, and 19 to 24 months. Household income data were missing for 61 participants and were analysed on an available-case basis. Because outcome classification was incomplete for some variables, underweight, wasting, and anaemia analyses were also conducted on an available-case basis. Associations between nutritional status indicators and categorical variables were examined using chi-square tests, or Fisher’s exact test where chi-square assumptions were not met. Pearson correlation was used to assess the relationship between knowledge and practice scores. Independent-samples t-tests were used to compare mean knowledge and practice scores across nutritional status categories. Statistical significance was set at *p*<0.05.

In secondary analyses, parsimonious multivariable logistic regression models were fitted for stunting and underweight using child age group (6-12 vs 13-24 months), sex, previous malnutrition treatment, and either a five-item household adversity index or household food shortage, with maternal knowledge and practice scores entered in separate models. Given the limited number of outcome events, these adjusted models were intended as exploratory parsimonious analyses to examine broad patterns of association rather than to support detailed causal inference. The household adversity index assigned one point each for war-related displacement, household food shortage, receipt of food aid, non-municipal drinking water, and paternal unemployment. Because only 11 children were classified as wasted, no multivariable wasting model was fitted. Secondary linear regression models examined associations of PHCC service variables with maternal knowledge and practice scores.

## Results

Among 200 children analysed, 25 (12.5%) were classified as stunted. Underweight classification was available for 199 children, of whom 40 (20.1%) were classified as underweight, and wasting classification was available for 53 children, of whom 11 (20.8%) were classified as wasted. Among the 55 children with available haemoglobin data, 35 (63.6%) had anaemia. Table 1 summarises anthropometric and anaemia findings within the haemoglobin-tested subsample. Study recruitment, exclusions, and derivation of the haemoglobin-tested subsample are summarised in Figure 1.

**Table 1.** Anthropometric outcomes and anaemia findings in the haemoglobin-tested subsample.

| Indicator | n | % |
| --- | --- | --- |
| Stunting (n=47)<sup>a</sup> | 4 | 8.5 |
| Wasting (n=11)<sup>b</sup> | 4 | 36.4 |
| Underweight (n=52)<sup>c</sup> | 11 | 21.2 |
| Anaemia (n=55)<sup>d</sup> | 35 | 63.6 |
| Mild anaemia among tested children (n=55)<sup>d</sup> | 5 | 9.1 |
| Moderate anaemia among tested children (n=55)<sup>d</sup> | 28 | 50.9 |
| Severe anaemia among tested children (n=55)<sup>d</sup> | 2 | 3.6 |

Note. Findings are presented for the **haemoglobin-tested subsample**. Haemoglobin was available for 55 children, and anaemia was defined from the numeric haemoglobin value as **Hb <11 g/dL**. Mild anaemia was defined as **10.0-10.9 g/dL**, moderate anaemia as **7.0-9.9 g/dL**, and severe anaemia as **<7.0 g/dL**.
**a** Stunting classification was available for 47 of the 55 haemoglobin-tested children.
**b** Wasting classification was available for 11 of the 55 haemoglobin-tested children.
**c** Underweight classification was available for 52 of the 55 haemoglobin-tested children.
**d** Percentages for anaemia and anaemia severity were calculated using 55 as the denominator.

More stunting cases were observed among younger infants, but stunting was not significantly associated with child age category (p=0.177). Because the analysis did not calculate age-specific prevalence within each age band, this finding should be interpreted as a distribution of cases rather than a direct estimate of age-specific risk. Stunting showed no significant associations with maternal education, paternal education, family size, displacement status, food shortage, or previous malnutrition treatment (all *p*>0.05).

Underweight was more common among children from households reporting food shortage than among those not reporting food shortage (33/131 [25.2%] vs 7/68 [10.3%], Pearson’s χ^2^=6.185, p=0.013). Underweight was also more common among children with previous malnutrition treatment than among those without previous treatment (23/73 [31.5%] vs 17/126 [13.5%], Pearson’s χ^2^=9.340, p=0.002) (Table 2).

**Table 2.** Significant Associations with Malnutrition Outcomes.

| Indicator | Determinant | Higher-risk group, n/N (%) | Comparison group, n/N (%) | Test | P-value |
| --- | --- | --- | --- | --- | --- |
| Underweight | Food shortage | With food shortage: 33/131 (25.2%) | Without food shortage: 7/68 (10.3%) | Pearson's $\chi^2 = 6.185$ | 0.013 |
| Underweight | Previous malnutrition Rx | Previous Rx: 23/73 (31.5%) | No previous Rx: 17/126 (13.5%) | Pearson's $\chi^2 = 9.340$ | 0.002 |
| Wasting □ | Child age category | Peak at 13-18 months: 5/9 (55.6%) | Lowest at 7-9 months: 0/13 (0.0%) | Fisher's exact test | 0.0024 |
| Anaemia □ | Paternal employment | Father not employed: 31/43 (72.1%) | Father employed: 4/12 (33.3%) | Fisher's exact test | 0.020 |

Note. Rx = treatment. <sup>a</sup> Based on the 51 children with both an available wasting classification and age in the analysed 6 to 24 month categories. <sup>b</sup> Based on the 55 haemoglobin-tested children, with anaemia defined from the numeric haemoglobin value as **Hb <11 g/dL**.

Wasting varied significantly across age categories in the subset with both wasting classification and age available (Fisher’s exact test, p=0.0024). The highest wasting proportion was observed in the 13–18-month category (5/9, 55.6%), whereas no wasting cases were observed in the 7–9-month category (0/13). Because only 10 wasted children had age data in this subset, this finding should be interpreted cautiously.

Among the 55 children with available haemoglobin data, paternal employment was significantly associated with anaemia (Fisher’s exact test, p=0.020), with anaemia more frequent among children whose fathers were not employed than among those whose fathers were employed (31/43 [72.1%] vs 4/12 [33.3%], p=0.020). No significant associations were found with child age, maternal education, family size, displacement, food aid receipt, or previous malnutrition treatment. The small, non-random haemoglobin-tested subsample constrains the generalisability of these findings. Complete bivariate analyses, including non-significant associations, are presented in Supplementary Tables S2-S17 (stunting:S2-S5; underweight:S6-S9; wasting:S10-S13; anaemia:S14-S17).

Regarding healthcare service delivery, 78.0% reported regular PHCC attendance, and 71.0% of mothers reported that PHCC nurses provided nutrition advice. In a separate item on guidance type, counselling content included breastfeeding guidance (50.5%), complementary feeding advice (28.0%), food hygiene (16.0%), and anaemia or supplementation information (11.5%); 21.0% selected ‘no guidance was provided at all’. Only 52.5% reported growth-chart review with nurses. These counselling-content percentages were based on the guidance-type item and were not mutually exclusive. Detailed item-level results for child feeding practices and PHCC nutrition service delivery are provided in Supplementary Tables S18 and S19, respectively.

Maternal knowledge scores averaged 9.5±1.4 (observed range 5–11), indicating generally high knowledge. Specific gaps: only 67.5% correctly understood exclusive breastfeeding definition; 27.5% incorrectly believed breastfeeding should reduce during illness. Practice scores averaged 51.5 ± 7.5 (observed range 20-60). Important practice gaps remained: only 52.5% reported always practising exclusive breastfeeding until 6 months, 61.0% reported always avoiding sugary drinks, and 57.5% reported always limiting added sugar/salt. Item-level distributions for maternal knowledge and practice, together with descriptive score summaries, are shown in Supplementary Tables S20-S22.

The retained knowledge and practice scales showed acceptable internal consistency (Cronbach’s alpha 0.740 and 0.780, respectively), and knowledge and practice scores were positively, although modestly, correlated (r=0.177, p=0.012). The full knowledge-practice correlation matrix is presented in Supplementary Table S23. Knowledge scores were not significantly associated with stunting (p=0.324), underweight (p=0.873), wasting (p=0.127), or anaemia (p=0.654) (Supplementary Table S24). Practice scores were not significantly associated with stunting (p=0.124), underweight (p=0.753), wasting (p=0.527), or anaemia (p=0.702) (Supplementary Table S25).

Adjusted models for stunting and underweight are summarised in Table 3, with full model outputs presented in Supplementary Table S26. In adjusted logistic regression analyses, younger age was associated with higher odds of stunting, reaching statistical significance in the knowledge model (aOR 2.93, 95% CI 1.01 to 8.49, p=0.048) and borderline significance in the practice model (aOR 2.88, 95% CI 1.00 to 8.30, p=0.050). For underweight, previous malnutrition treatment remained independently associated with higher odds of underweight (aOR 3.56, 95% CI 1.61 to 7.87, p=0.002 in the knowledge model; aOR 3.49, 95% CI 1.59 to 7.66, p=0.002 in the practice model). Maternal knowledge and practice scores were not independently associated with stunting or underweight in adjusted models, and the five-item household adversity index was not independently associated with either outcome. In secondary pathway-oriented analyses, receipt of written or visual educational materials was associated with a modestly higher maternal knowledge score (β 0.45, 95% CI 0.02 to 0.89, p=0.040), and higher maternal knowledge score was associated with a higher maternal practice score (β 0.86, 95% CI 0.13 to 1.60, p=0.022) (Supplementary Table S27). In sensitivity analyses replacing the household adversity index with household food shortage, the overall pattern was similar: younger age remained associated with stunting, previous malnutrition treatment remained associated with underweight, and household food shortage was attenuated after adjustment (Supplementary Table S28).

**Table 3.**
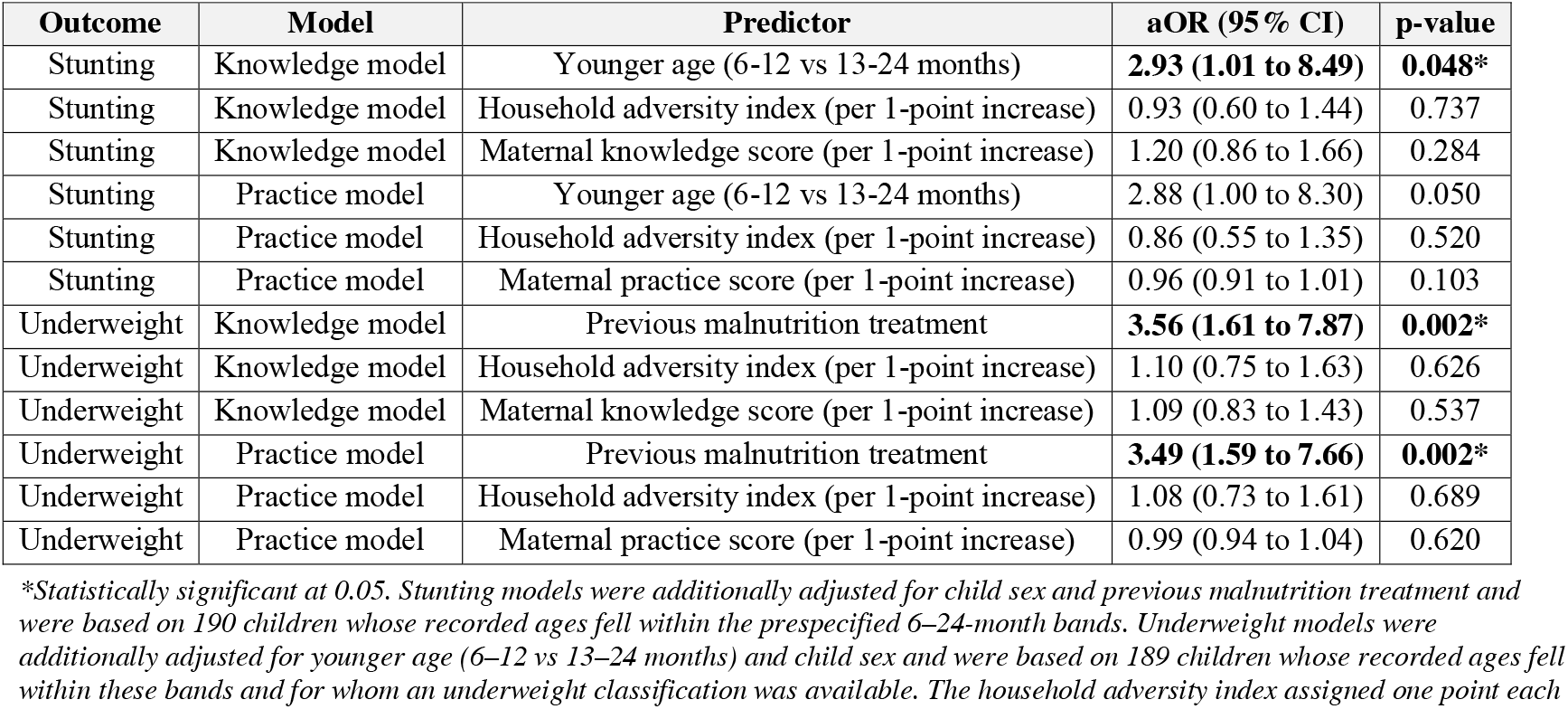
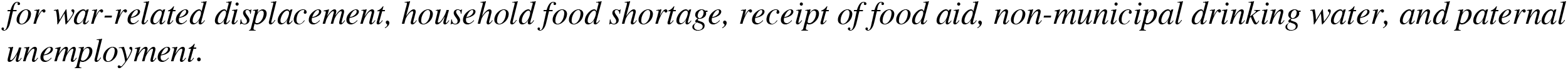
Adjusted logistic regression models for stunting and underweight.

| Outcome | Model | Predictor | aOR (95% CI) | p-value |
| --- | --- | --- | --- | --- |
| Stunting | Knowledge model | Younger age (6-12 vs 13-24 months) | **2.93 (1.01 to 8.49)** | **0.048\*** |
| Stunting | Knowledge model | Household adversity index (per 1-point increase) | 0.93 (0.60 to 1.44) | 0.737 |
| Stunting | Knowledge model | Maternal knowledge score (per 1-point increase) | 1.20 (0.86 to 1.66) | 0.284 |
| Stunting | Practice model | Younger age (6-12 vs 13-24 months) | 2.88 (1.00 to 8.30) | 0.050 |
| Stunting | Practice model | Household adversity index (per 1-point increase) | 0.86 (0.55 to 1.35) | 0.520 |
| Stunting | Practice model | Maternal practice score (per 1-point increase) | 0.96 (0.91 to 1.01) | 0.103 |
| Underweight | Knowledge model | Previous malnutrition treatment | **3.56 (1.61 to 7.87)** | **0.002\*** |
| Underweight | Knowledge model | Household adversity index (per 1-point increase) | 1.10 (0.75 to 1.63) | 0.626 |
| Underweight | Knowledge model | Maternal knowledge score (per 1-point increase) | 1.09 (0.83 to 1.43) | 0.537 |
| Underweight | Practice model | Previous malnutrition treatment | **3.49 (1.59 to 7.66)** | **0.002\*** |
| Underweight | Practice model | Household adversity index (per 1-point increase) | 1.08 (0.73 to 1.61) | 0.689 |
| Underweight | Practice model | Maternal practice score (per 1-point increase) | 0.99 (0.94 to 1.04) | 0.620 |

\*Statistically significant at 0.05. Stunting models were additionally adjusted for child sex and previous malnutrition treatment and were based on 190 children whose recorded ages fell within the prespecified 6–24-month bands. Underweight models were additionally adjusted for younger age (6–12 vs 13–24 months) and child sex and were based on 189 children whose recorded ages fell within these bands and for whom an underweight classification was available. The household adversity index assigned one point each
*for war-related displacement, household food shortage, receipt of food aid, non-municipal drinking water, and paternal unemployment.*

## Discussion

This study provides rare clinic-based evidence from active conflict conditions in Gaza from Al-Daraj Martyrs Health Centre, one of the remaining functioning PHCCs in Gaza City during the study period, showing that malnutrition among children aged 6-24 months was not only common but embedded within simultaneous food-system collapse and erosion of PHCC nutrition services. Its distinctive contribution lies not simply in documenting undernutrition in a hard-to-study humanitarian setting, but in linking child nutritional status with household deprivation, service erosion and maternal domains within the same analytic framework. Taken together, the findings suggest that the observed pattern is more consistent with structural deprivation than with maternal knowledge scores alone (8,14,16). In that sense, the study contributes not only prevalence data, but also sentinel operational evidence from the remaining routine PHCC access point for young children, showing that PHCC-based assessment can serve as a practical warning signal for nutritional deterioration when representative surveillance is difficult to sustain during active conflict (14,16).

Among the 55 children with available haemoglobin data, 35 (63.6%) had anaemia. This proportion was higher than the pre-conflict estimate of 35.6% reported among children aged 24-59 months in Gaza, but comparison should be cautious because the present estimate derives from a small, non-random haemoglobin-tested subsample rather than systematic screening (9). The finding is therefore best interpreted as a signal of concern rather than a population-level prevalence estimate. Within the haemoglobin-tested subsample, anaemia was more frequent among children whose fathers were not employed than among those whose fathers were employed (p=0.020), which may reflect wider household deprivation rather than a specific biological pathway. Anaemia in this setting is unlikely to reflect iron deficiency alone. In young children, it often arises from overlapping contributions including iron and other micronutrient deficiencies, infection or inflammation, and inherited red-cell disorders. In Gaza, where dietary quality, supplementation coverage, and infection control have all deteriorated, a multifactorial explanation is especially plausible (8,14,16,26).

Underweight was more common among children from households reporting food shortage than among those not reporting food shortage (p=0.013), linking household deprivation directly to child undernutrition in this sample and aligning with recent reports of marked dietary deterioration in Gaza (8). Similar associations between household food insecurity and childhood undernutrition have been reported in other settings (27).

The association between previous malnutrition treatment and current underweight (p=0.002) raises programmatic concern and may reflect relapse after treatment, interruption of follow-up, or persistent underlying risk factors. Systematic review evidence indicates that relapse after treatment for severe acute malnutrition remains a recognised problem even in stable settings (28), and may be even more difficult to prevent under conflict conditions. In addition, barriers to timely care-seeking, reaching services, and receiving adequate care may all reduce effective treatment coverage, which is especially relevant where malnutrition services operate under conflict-disrupted conditions (29). Adjusted analyses refined the bivariate picture. Younger age remained independently associated with stunting, and previous malnutrition treatment remained independently associated with underweight. By contrast, maternal knowledge score, maternal practice score, and the cumulative household adversity index were not independently associated with either outcome. In sensitivity analyses replacing the adversity index with household food shortage, the broad pattern was unchanged, but the food-shortage association with underweight attenuated after adjustment, suggesting that the bivariate relationship may reflect wider household vulnerability rather than a single independent exposure.

Although more stunting cases were observed among younger infants, this did not show a statistically significant association between stunting and child age category, and the study did not estimate age-specific prevalence within each age band. Although lower than the underweight burden observed here, this level of stunting remains important because stunting is a core global nutrition target and is associated with long-term developmental risk (4,5,30). Early stunting often reflects fetal growth restriction and/or immediate postnatal inadequacy (5,31), and evidence from very young infants also underscores how severe nutritional vulnerability can emerge early in life when maternal and early postnatal risks accumulate (32). In this context, the pattern may point to maternal undernutrition during pregnancy and early infancy. The lack of socioeconomic associations contrasts with peacetime patterns where strong gradients consistently emerge, with maternal, environmental, and household factors all contributing importantly to stunting risk (33,34,35), potentially reflecting a crisis-related compression of risk in which traditional protective factors are eroded.

Wasting classification was available for 53 children, of whom 11 (20.8%) were classified as wasted. This available-case estimate is not directly comparable with recent Gaza surveillance based on MUAC-defined acute malnutrition, but together the findings suggest substantial acute nutritional vulnerability among children aged 6 to <24 months (13). Recent pooled longitudinal analyses from low- and middle-income countries also show that wasting and stunting frequently cluster in the same highly vulnerable early-life period (36). Evidence from children with severe acute malnutrition also suggests that concurrent anthropometric failure is associated with overlapping determinants such as younger age, poverty, illness, poor sanitation, and limited healthcare access (37). This comparison should be interpreted cautiously, because the present study used weight-for-height Z-scores whereas the cited Gaza surveillance used MUAC-based case definitions. These indicators identify overlapping but non-identical groups of children, so apparent discrepancies do not necessarily indicate a true epidemiological contradiction (38,39). This discrepancy likely reflects the available-case nature of the wasting analysis, the small number of wasted children, PHCC-attending children representing a healthier subset, and measurement timing variability. The observed clustering of wasting cases in the 13-18-month category is consistent with the recognised vulnerability of children during the complementary-feeding period (2,40), but this should be interpreted cautiously because the study did not estimate age-specific prevalence within each age band.

Analysis of PHCC service delivery revealed important gaps. Nearly one-third of mothers reported not receiving nutritional advice from nurses, 21.0% reported that no guidance was provided on the separate guidance-type item, only 52.5% had growth charts reviewed, and complementary feeding advice reached just 28.0%. These deficiencies are more likely to reflect erosion of system capacity than individual provider performance (14,16). That interpretation is reinforced by the fact that the study site was one of the remaining functioning PHCCs in Gaza City during the study period, suggesting broader system compression rather than an isolated problem at one centre. This matters because growth monitoring and promotion is intended to combine regular child measurement with counselling and follow-up, yet review evidence indicates that its impact is limited and uncertain when promotion activities are weak or inconsistently delivered (41). At the same time, locally adapted breastfeeding-support initiatives in Gaza have shown that health-worker training, remote mentorship and mother-focused lactation support can still be delivered despite severe disruption (42). Together with wider accounts of conflict-related damage to food systems and health services in Gaza (43), these findings suggest that the problem is not only reduced access to care, but also reduced capacity to deliver meaningful nutrition support. Because the sample included only families able to reach this PHCC, the findings should be interpreted as sentinel evidence from a remaining access point rather than as a full picture of Gaza (14,16).

Maternal knowledge and practice scores were relatively high, the retained scales showed acceptable internal consistency, and the two scores were weakly but significantly correlated (r=0.177, p=0.012). Under conditions of severe food scarcity and service disruption, however, maternal knowledge may not translate reliably into protective feeding practices because household agency is profoundly constrained (8,14,16). In this sample, practice scores were not associated with child nutritional outcomes, and knowledge scores were not significantly associated with stunting, underweight, wasting, or anaemia. The wasting comparison was based on a smaller subset with available wasting classification and should therefore be interpreted cautiously. This should not be interpreted as evidence that feeding behaviour is unimportant. Rather, it suggests that in a crisis setting, caregiver knowledge alone may be insufficient to offset structural deprivation.

Secondary pathway-oriented analyses suggested a limited but coherent pattern within the maternal domains. Receipt of written or visual educational materials was associated with slightly higher maternal knowledge, and higher knowledge was associated with better reported practice. However, these score-based improvements did not translate into independent associations with stunting or underweight in adjusted models. This pattern supports the interpretation that, in this setting, education and counselling may improve knowledge and reported practice, but are unlikely to offset severe constraints on food access and continuity of care.

The feeding data reinforce the interpretation that structural deprivation, rather than knowledge deficits alone, was central to nutritional risk in this sample. Consumption of meat or legumes was extremely limited, with only 7.0% of children reported to have consumed these foods in the previous 24 hours, whereas 54.0% consumed dairy and 53.5% received micronutrient supplements. These patterns suggest constrained dietary availability and poor micronutrient access, which may help explain why relatively high maternal knowledge scores did not translate into better anthropometric outcomes.

Evidence from more stable settings is mixed. A Ghanaian cross-sectional study likewise found no significant association between maternal nutrition knowledge and child malnutrition status (20). By contrast, a Nigerian study reported poor complementary-feeding knowledge and practices, low dietary diversity, and low minimum acceptable diet, particularly among less educated mothers, supporting the importance of caregiver education for feeding quality in non-crisis settings (44). Reviews similarly suggest that nutrition education and caregiver feeding support can improve feeding practices, dietary quality, and, in some settings, child nutritional outcomes (15,21). In this context, counselling remains necessary, but is unlikely to provide measurable nutritional protection without concurrent food access, micronutrient provision, and continuity of care, consistent with recent WHO guidance on wasting management and complementary feeding (45,40). In addition, because practice data were self-reported in face-to-face interviews, social desirability bias may have reduced variation in reported practices and attenuated practice-outcome associations (46).

This finding carries important intervention implications: conventional nutrition education approaches, while valuable in stable contexts, are unlikely to be sufficient under catastrophic conditions. These findings support prioritising structural interventions, including emergency food assistance and restoration of healthcare capacity (8,14,43,16). This interpretation is also consistent with recent WHO guidance, which emphasises integrated prevention and management of wasting within functioning health systems, alongside age-specific complementary-feeding support for children aged 6-23 months. In Gaza, counselling should therefore be preserved, but it cannot substitute for food access, micronutrient provision, referral pathways, and basic service restoration (45,40).

This study has a few limitations that may constrain interpretation. First, convenience sampling from a single PHCC introduces selection bias. However, this site was one of the remaining functioning PHCCs in Gaza City during the study period, which gives the findings particular sentinel value as evidence from the remaining routine primary-care entry point for young children. Even so, the results should not be interpreted as population-representative estimates for Gaza as a whole, because they exclude children who could not reach this service. Second, 50 questionnaires were excluded because anthropometric records were incomplete, which may have biased the analytic sample towards children with better documentation. Third, cross-sectional design precludes causal inference, and the adjusted logistic regression analyses should be interpreted as exploratory because the limited number of stunting and underweight events constrained model complexity and may have increased the risk of overfitting. Fourth, anaemia assessment was available in only 27.5% of the sample (55/200) and was based on a selective haemoglobin-tested subsample, which severely limits generalisability and prevents interpretation as a cohort-wide prevalence estimate. Fifth, reliance on medical records prevents measurement quality verification. Sixth, data collection during active conflict likely introduced systematic errors. Finally, household income had substantial missingness, which limited interpretation of income-related associations. Despite these limitations, the findings support an urgent emergency response that should include micronutrient supplementation, expansion of therapeutic feeding programmes, emergency food assistance for families with young children, strengthening of PHCC capacity, and support for maternal nutrition.

In this single-centre, clinic-based sample from one of the remaining functioning PHCCs in Gaza City during the study period, children aged 6-24 months experienced a substantial burden of malnutrition, and the findings provide sentinel evidence of how structural deprivation and PHCC service erosion were shaping nutritional risk in early life. Anaemia was also frequent among the haemoglobin-tested subsample. The pattern of associations suggests that conflict-related structural deprivation, especially household food shortage and disrupted nutrition services, may be more closely aligned with nutritional risk than maternal knowledge alone in this clinic-based sample. In adjusted analyses, younger age was associated with stunting and previous malnutrition treatment was associated with underweight, whereas maternal knowledge and practice scores were not independently associated with anthropometric outcomes. These findings support integrated responses that combine food access, micronutrient provision, follow-up of previously malnourished children, and restoration of core PHCC nutrition functions.

## Data Availability Statement

De-identified participant data available from corresponding author upon reasonable request, subject to ethical approval and data protection regulations.

## Acknowledgements

For the purpose of open access, the author has applied a Creative Commons Attribution (CC BY) licence to any Author Accepted Manuscript version arising from this submission.

## Author Contributions

LM: data collection, analysis and interpretation. HA and AN: study conception and design and revised the manuscript for intellectual content. JR drafted the manuscript, critically revised the manuscript and contributed to data interpretation. RA: contributed to statistical analysis, data interpretation and critically revised the clinical aspects of manuscript. KM provided senior oversight, led manuscript development, contributed to the interpretation of data and critically revised it for scientific rigour. All authors approved the final version of the manuscript and agree to be accountable for all aspects of the work.

## Ethical Approval

Ethical approval was obtained prior to data collection from the Ministry of Health, State of Palestine, Human Resources Development Directorate (reference number 25121016; 25 June 2025) and from Al-Quds University (24 August 2025). Written informed consent was obtained from all participants.

