## Supplemental Files for "Nutritional Status and Associated Factors Among Children Aged 6-24 Months at a Primary Health Care Centre in Conflict-Affected Gaza"

### Supplementary Information

**Questionnaires:** English version

#### Domain 1: Child Nutritional Status

| No. | Question | Answer |
|---|---|---|
| NS 01 | Child's age (in months) | ________________ |
| NS 02 | Child's weight | ________________ |
| NS 03 | Child's length | ________________ |
| NS 04 | Any chronic condition affecting nutrition? | ❑ Yes<br>❑ No If yes, specify/ |
| NS 05 | Hb value (from available laboratory result in the child's health file) | ____________________ |
| NS 06 | Does the child have anaemia? | ❑ Yes /<br>❑ No / (If no, skip next question) |
| NS 07 | If yes, what is the anaemia classification? | ❑ Mild / Moderate / Severe / |
| NS 08 | Has the child been treated for malnutrition previously? | ❑ Yes<br>❑ No<br>If yes, what treatment and for what |

#### Domain 2: Feeding Practices / المجال الثاني: ممارسات التغذية

| No. | Questions | Answer |
|---|---|---|
| FP 01 | Was the child breastfed? | ❑ Yes<br>❑ No |
| FP 02 | Exclusive breastfeeding duration (months) | ____________________________ |
| FP 03 | Feeding method | ❑ Breastfeeding only ❑ Bottle only ❑ Mixed |
| FP 04 | Has complementary feeding started? | ❑ Yes<br>❑ No |
| FP 05 | Age started (months) | ____________________________ |
| FP 06 | First type of food introduced | ❑ Juice / ❑ Cereals /<br>❑ Vegetables / ❑ Fruits / ❑ Other /<br>__________ |
| FP 07 | Has breastfeeding stopped? | ❑ Yes<br>❑ No (If no, skip next question) |
| FP 08 | Age of stopping | ____________________________ |
| FP 09 | Number of meals per day | ____________________________ |
| FP 010 | Food groups consumed in the last 24 hours | ❑ cereals/ ❑ Vegetables / ❑ Fruits /<br>❑ Meat & legumes/ ❑ Dairy / ❑ Sweets and oils / |
| FP 011 | Do you add micronutrient supplements? | ❑Yes<br>❑ No |
| FP 012 | Feeding difficulties in the last week? | ❑ Yes (if yes, specify and for how many days ?)<br>❑ No |

**Domain 3: Socio-demographic, Economic & Environmental Factors**

| No. | Question | Answer |
|---|---|---|
| SD 01 | Mother's age | ___________________________ |
| SD 02 | Mother's education | ❑ Illiterate<br>❑ Primary<br>❑ Preparatory<br>❑ Secondary<br>❑ University |
| SD 03 | Father's education | ❑ Illiterate<br>❑ Primary<br>❑ Preparatory<br>❑ Secondary<br>❑ University |
| SD 04 | Number of children in the child's family | ___________________________ |
| SD 05 | Child's birth order | ___________________________ |
| SD 06 | Child's sex | ❑ Male ❑ Female |
| SD 07 | Father's employment status | ❑ Yes ❑ No |
| SD 08 | Mother's employment status | ❑ Yes ❑ No |
| SD 09 | Monthly household income | NIS |
| SD 010 | Was the family displaced due to war? | ❑ Yes<br>❑ No (If no, skip the next question) |
| SD 011 | If yes, displacement duration | Months |
| SD 012 | Does the family face food shortages? | ❑ Yes<br>❑ No (If no, skip next question) |
| SD 013 | If yes, the food shortage duration | Months |
| SD 014 | Does the family receive food aid? | ❑ Yes ❑ No |
| SD 015 | Source of drinking water | ❑ Municipal<br>❑ Tanker<br>❑ Bottled<br>❑ Other ___________ |

**Domain 4: Health Services & Nurses' Role**

| No. | Question | Answer |
|---|---|---|
| HSNR 01 | Do you visit the child health center regularly? | ❑ Yes<br>❑ No |
| HSNR 02 | If not, why? | |
| HSNR 03 | Do nurses at PHC centers provide nutrition advice? | ❑ Yes<br>❑ No |
| HSNR 04 | Type of guidance provided | ❑ Breastfeeding<br>❑ Complementary feeding<br>❑ Food hygiene<br>❑ Anemia & supplements<br>❑ No advice |

| | | |
|---|---|---|
| HSNR 05 | Do you believe nurses play a role in improving your child's nutrition? | ❑Yes<br>❑ No |
| HSNR 06 | Did the nurse assess your child's growth charts with you? | ❑Yes<br>❑ No |
| HSNR 07 | Did you receive written or visual educational materials? | ❑Yes<br>❑ No |
| HSNR 08 | Suggestions to improve nurses' roles | |

**Domain 5: Mothers' Knowledge on Breastfeeding & Complementary /Feeding**

| Question No. | Statements | Answer |
|---|---|---|
| MK 01 | Breastfeeding should start within the first hour after birth. | ☐Yes<br>☐ No |
| MK 02 | Colostrum is beneficial and should be given to the infant. | ☐Yes<br>☐ No |
| MK 03 | Exclusive breastfeeding means giving only breast milk without water or herbs until 6 months. | ☐Yes<br>☐ No |
| MK 04 | Pacifier use early on does not affect breastfeeding. | ☐Yes<br>☐ No |
| MK 05 | Promising signs of milk adequacy include sufficient urination and weight gain. | ☐Yes<br>☐ No |
| MK 06 | It is preferable to reduce breastfeeding when the infant is ill. | ☐ Yes<br>☐ No |
| MK 07 | Correct positioning and latch reduce nipple cracks and engorgement. | ☐Yes<br>☐ No |
| MK 08 | Breastfeeding alongside food is recommended until 24 months or more. | ☐ Yes<br>☐ No |
| MK 09 | Complementary feeding should start at 6 months with continued breastfeeding. | ☐ Yes<br>☐ No |
| MK 010 | Food consistency should progress from smooth purees to more solid textures as the child ages. | ☐ Yes<br>☐ No |
| MK 011 | Iron-rich/fortified foods are essential to prevent anaemia. | ☐ Yes<br>☐ No |
| MK 012 | Sugar/salt should be avoided in infant food. | ☐ Yes<br>☐ No |
| MK 013 | Food hygiene and safety are not important at this age. | ☐ Yes<br>☐ No |
| MK 014 | Sugary drinks and energy drinks are suitable for infants. | ☐ Yes<br>☐ No |

**Domain 6: Mothers' practice on Breastfeeding & Complementary /Feeding**

| Question No. | Statements | Always | Often | Sometimes | Rarely | Never |
|---|---|---|---|---|---|---|
| MP 01 | I started breastfeeding my baby within the first hour after birth. | | | | | |

| | |
|---|---|
| MP 02 | I gave my baby colostrum and did not discard it. |
| MP 03 | I practised exclusive breastfeeding until 6 months without water/herbs/formula. |
| MP 04 | I used a pacifier with my baby during the first six months. |
| MP 05 | I increase or maintain breastfeeding frequency when my baby is ill. |
| MP 06 | I check my baby's positioning and latch during breastfeeding. |
| MP 07 | I continued breastfeeding alongside complementary feeding after 12 months. |
| MP 08 | I started complementary feeding for my baby at 6 months. |
| MP 09 | I gradually increase my child's food consistency as they get older. |
| MP 010 | I regularly provide iron-rich or fortified foods. |
| MP 011 | I avoid adding sugar/salt to my child's food or keep them to a minimum. |
| MP 012 | I wash my hands with safe water and use clean utensils when preparing/feeding the child. |
| MP 013 | I avoid giving sugary/soft/energy drinks to my child. |

**Note: The full questionnaire included 14 knowledge items and 13 practice items. After internal consistency testing, three knowledge items and one practice item were excluded from the final composite scoring. Therefore, the reliability analysis and score-based results reported in Supplementary Tables S1 and S20–S25 are based on 11 knowledge items and 12 practice items.*

**Questionnaires:** Arabic version

| المجال الأول: الحالة التغذوية للطفل | | |
|---|---|---|
| الرقم | السؤال | الإجابة |
| 1. | عمر الطفل | ________________شهر |
| 2. | وزن الطفل | ________________كغم |
| 3. | طول الطفل | ________________سم |
| 4. | هل يعاني الطفل من أمراض مزمنة تؤثر على تغذيته؟ | ☐ نعم |
| | | ☐ لا |
|  |  | لو الإجابة نعم حددي---------------------- |
| 5. | نسبة الهيموغلوبين (حسب ملف الطفل) | …………………………………………….. |

| | | |
|---|---|---|
| 6. | هل يعاني الطفل من فقر الدم؟ | ☐ نعم<br>☐ لا<br>اذا لا تخطى السؤال التالي |
| 7. | إذا نعم، ما هو تصنيف فقر الدم؟ | ☐ خفيف<br>☐ متوسط<br>☐ شديد |
| 8. | هل تم علاج الطفل من سوء التغذية سابقًا؟ | ☐ نعم<br>☐ لا<br>اذا كانت الإجابة نعم اين وما هي المشكلة والعلاج المقدم ----------------- |
| **المجال الثاني: ممارسات التغذية** | | |
| FP 01 | هل تم إرضاع الطفل طبيعيًا؟ | ☐ نعم<br>☐ لا |
| FP 02 | مدة الرضاعة وحدها قبل ادخال أطعمة أخرى للطفل (بالأشهر) | ________________ |
| FP 03 | طريقة التغذية | ☐ رضاعة فقط<br>☐ رضاعة بالزجاجة فقط<br>☐ مختلطة |
| FP 04 | هل بدأ الطفل تغذية تكميلية؟ | ☐ نعم<br>☐ لا |
| FP 05 | العمر عند بداية التغذية التكميلية | ________________ |
| FP 06 | نوع الغذاء الأول | ☐ عصير<br>☐ حبوب<br>☐ خضروات<br>☐ فواكه<br>☐ أخرى |
| FP 07 | هل توقفتِ عن الإرضاع الطبيعي؟ | ☐ نعم<br>☐ لا |

| | | |
|---|---|---|
| | | اذا كانت الإجابة لا تخطى السؤال التالي |
| FP 08 | العمر عند توقف الرضاعة الطبيعية | ____________________ |
| FP 09 | عدد الوجبات اليومية | ____________________ |
| FP 010 | المجموعات الغذائية خلال 24 ساعة | ☐ الحبوب |
| | | ☐ الخضراوات |
| | | ☐ الفواكه |
| | | ☐ بقوليات ولحوم |
| | | ☐ الحليب |
| | | ☐ الحلويات والزيوت |
| FP 011 | هل تضيفين مساحيق المغذيات الدقيقة أو المكملات؟ | ☐ نعم |
| | | ☐ لا |
| FP 012 | هل واجه الطفل صعوبات | ☐ نعم |
| | | ☐ لا |
| | | اذا كانت الإجابة نعم حددي وكم المدة؟ ......................................................... |
| **المجال الثالث: العوامل الاجتماعية والديموغرافية والاقتصادية والبيئية** | | |
| SD 01 | عمر الأم | ____________________ |
| SD 02 | تعليم الأم | ☐ لا تقرأ ولا تكتب |
| | | ☐ ابتدائي |
| | | ☐ إعدادي |
| | | ☐ ثانوي |
| | | ☐ جامعي |
| SD 03 | تعليم الأب | ☐ لا يقرأ ولا يكتب |
| | | ☐ ابتدائي |
| | | ☐ إعدادي |
| | | ☐ ثانوي |
| | | ☐ جامعي |
| SD 04 | عدد الأطفال في أسرة الطفل | ____________________ |
| SD 05 | ترتيب الطفل بين إخوته | ____________________ |
| SD 06 | جنس الطفل | ☐ ذكر |
| | | ☐ أنثى |
| SD 07 | هل الأب يعمل؟ | ☐ نعم |
| | | ☐ لا |
| SD 08 | هل الأم تعمل؟ | ☐ نعم |

| | | |
|---|---|---|
| | | ☐ لا |
| SD 09 | الدخل الشهري للأسرة | ___________ NIS/شيكل |
| SD 010 | هل تم نزوح الأسرة بسبب الحرب؟ | ☐ نعم<br>☐ لا<br>اذا كانت الإجابة " لا" تخطى السؤال التالي |
| SD 011 | إذا نعم، ما المدة لفترة النزوح؟ | ___________ شهرًا |
| SD 012 | هل تعاني الأسرة من نقص في الغذاء؟ | ☐ نعم<br>☐ لا<br>اذا كانت الإجابة " لا" تخطى السؤال التالي |
| SD 013 | إذا نعم، كم المدة التي عانت فيها الأسرة من نقص الغذاء؟ | ___________ شهرًا |
| SD 014 | هل تتلقى الأسرة مساعدات غذائية؟ | ☐ نعم<br>☐ لا |
| SD 015 | مصدر مياه الشرب | ☐ بلدية<br>☐ صهريج<br>☐ معبأة<br>☐ أخرى |
| **المجال الرابع: دور التمريض والخدمات الصحية** | | |
| HSNR 01 | هل ترددين بطفلك على مركز متابعة صحة الطفل بشكل دوري؟ | ☐ نعم<br>☐ لا |
| HSNR 02 | إذا كانت الإجابة لا، لماذا؟ | -------------------------------- |
| HSNR 03 | هل تقدم الممرضة نصائح تغذوية؟ | ☐ نعم<br>☐ لا |
| HSNR 04 | نوع التعليمات المقدمة | ☐ الرضاعة<br>☐ التغذية التكميلية<br>☐ نظافة الغذاء<br>☐ فقر الدم والمكملات<br>☐ لا يوجد |
| HSNR 05 | هل ترين أن للممرضة دورًا في تحسين تغذية طفلك؟ | ☐ نعم<br>☐ لا |
| HSNR 06 | هل قامت الممرضة بمراجعة مخططات نمو طفلك معك؟ | ☐ نعم<br>☐ لا |
| HSNR 07 | هل تلقيتِ مواد تعليمية مكتوبة أو مرئية؟ | ☐ نعم<br>☐ لا |
| HSNR 08 | اقتراحات لتحسين دور الممرضة | ------------------------------------------- |
| **المجال الخامس: معرفة الأمهات حول الرضاعة الطبيعية والتغذية التكميلية** | | |
| MK 01 | يجب بدء الرضاعة الطبيعية خلال الساعة الأولى بعد الولادة | ☐ نعم<br>☐ لا |

| | | |
|---|---|---|
| MK 02 | اللبأ (الحليب الأول مفيد ويجب إعطاؤه للرضيع) | ☐ نعم<br>☐ لا |
| MK 03 | الرضاعة الطبيعية الحصرية تعني إعطاء حليب الأم فقط دون ماء أو أعشاب حتى عمر 6 أشهر. | ☐ نعم<br>☐ لا |
| MK 04 | يمكن استخدام اللهاية (المصاصة) مبكرًا دون أن تؤثر على الرضاعة. | ☐ نعم<br>☐ لا |
| MK 05 | العلامات الجيدة لكفاية الحليب تشمل تبوّل الطفل مرات كافية وزيادة وزنه. | ☐ نعم<br>☐ لا |
| MK 06 | في حال مرض الرضيع يُفضّل تقليل الرضاعة الطبيعية. | ☐ نعم<br>☐ لا |
| MK 07 | الصحيحان يقللان من تشققات (Latch) الوضعية والتقفي الحلمة والاحتقان | ☐ نعم<br>☐ لا |
| MK 08 | يُنصح بالاستمرار في الرضاعة الطبيعية بجانب الطعام حتى عمر 24 شهرًا أو أكثر. | ☐ نعم<br>☐ لا |
| MK 09 | يجب بدء التغذية التكميلية عند عمر 6 أشهر مع الاستمرار بالرضاعة | ☐ نعم<br>☐ لا |
| MK 10 | قوام الطعام ينبغي أن يتدرج من مهروس ناعم إلى أكثر تماسكًا مع العمر | ☐ نعم<br>☐ لا |
| MK 11 | الأغذية الغنية بالحديد/المدعّمة ضرورية للوقاية من الأنيميا | ☐ نعم<br>☐ لا |
| MK 12 | ينبغي تجنّب إضافة السكر/الملح بكثرة في طعام الرضّع | ☐ نعم<br>☐ لا |
| MK 13 | النظافة وسلامة الغذاء غير مهمة في هذا العمر | ☐ نعم<br>☐ لا |
| MK 14 | المشروبات المُحلّاة ومشروبات الطاقة مناسبة للرضّع | ☐ نعم<br>☐ لا |

**المجال السادس: ممارسة الأمهات حول الرضاعة الطبيعية والتغذية التكميلية**

| | الفقرات | دائماً | غالباً | أحياناً | نادراً | أبداً |
|---|---|---|---|---|---|---|

| | | | | | | |
|---|---|---|---|---|---|---|
| | | | | | بدأتُ إرضاع طفلي خلال الساعة الأولى بعد الولادة | MP1 |
| | | | | | أعطيتُ طفلي اللبأ ولم أتخلص منه | MP2 |
| | | | | | لتزمتُ بالرضاعة الطبيعية الحصرية حتى 6 أشهر دون ماء/أعشاب/حليب صناعي | MP3 |
| | | | | | استخدمتُ اللهاية مع طفلي خلال الأشهر الستة الأولى | MP4 |
| | | | | | أزيد أو أحافظ على وتيرة الرضاعة عندما يمرض طفلي | MP5 |
| | | | | | أتحقق من وضعية طفلي والتقفي الصحيح خلال الرضاعة | MP6 |
| | | | | | واصلتُ الرضاعة مع التغذية التكميلية بعد عمر 12 شهرًا | MP7 |
| | | | | | بدأتُ إطعام طفلي أطعمة تكميلية عند تمام 6 أشهر | MP8 |
| | | | | | أزيد قوام الطعام تدريجيًا بما يلائم عمر طفلي | MP9 |
| | | | | | أقدّم مصادر غنية بالحديد أو أطعمة مدعّمة بالحديد بانتظام | MP10 |
| | | | | | لا أضيف السكر/الملح إلى أطعمة طفلي أو أبقِيهما في حدود قليلة جدًا | MP11 |
| | | | | | أغسل يديّ وأستخدم ماءً مأمونًا وأدوات نظيفة عند تحضير/إطعام الطفل | MP12 |
| | | | | | أتجنب إعطاء مشروبات مُحلّاة/غازية/طاقة لطفلي | MP13 |

**Supplementary Table S1.** Reliability of the knowledge and practice domains

| Domain | Number of questions | Cronbach's alpha coefficient |
|---|---|---|
| Knowledge | 11 | 0.740 |
| Practice | 12 | 0.780 |
| Total | 23 | 0.776 |

Note. *The full questionnaire contained 14 knowledge items and 13 practice items. The Cronbach's alpha coefficients reported in this table were derived from the pilot-testing phase (n=25). Composite scoring in the main study used 11 knowledge items and 12 practice items after item reduction. The scored knowledge set retained items on colostrum, exclusive breastfeeding, milk adequacy, breastfeeding during illness, positioning/latch, breastfeeding to 24 months or more, timing of complementary feeding, texture progression, avoiding sugar/salt, food hygiene, and avoiding sugary or energy drinks. The scored practice set excluded the colostrum practice item.*

**Supplementary Table S2.** Relationship between stunting and socio-economic factors (n=200)

| Sociodemographic factors | Normal n (%) | Stunting n (%) | Total | Test | P value |
|---|---|---|---|---|---|
| **Mothers' education** | | | | Fisher's exact test | 1.000 |
| Preparatory or less | 26 (14.9) | 3 (12.0) | 29 (14.5) | | |
| Secondary or more | 149 (85.1) | 22 (88.0) | 171 (85.5) | | |
| **Fathers' education** | | | | Pearson's $\chi^2$ = 0.084 | 0.772 |
| Preparatory or less | 54 (30.9) | 7 (28.0) | 61 (30.5) | | |
| Secondary or more | 121 (69.1) | 18 (72.0) | 139 (69.5) | | |
| **Number of children in the child's family** | | | | Fisher's exact test | 0.391 |
| One child | 28 (16.0) | 6 (24.0) | 34 (17.0) | | |
| 2 children or more | 147 (84.0) | 19 (76.0) | 166 (83.0) | | |

| | | | | | |
|---|---|---|---|---|---|
| **Child sex** | | | | Pearson's $\chi^2$ = 0.046 | 0.830 |
| Male | 95 (54.3) | 13 (52.0) | 108 (54.0) | | |
| Female | 80 (45.7) | 12 (48.0) | 92 (46.0) | | |
| **Father's employment status** | | | | Fisher's exact test | 1.000 |
| Yes | 34 (19.4) | 5 (20.0) | 39 (19.5) | | |
| No | 141 (80.6) | 20 (80.0) | 161 (80.5) | | |
| **Mothers' employment status** | | | | Fisher's exact test | 0.416 |
| Yes | 3 (1.7) | 1 (4.0) | 4 (2.0) | | |
| No | 172 (98.3) | 24 (96.0) | 196 (98.0) | | |
| **Income (n = 139)** | | | | Fisher's exact test | 1.000 |
| less than 1000 NIS | 111 (93.3) | 19 (95.0) | 130 (93.5) | | |
| 1000 NIS or more | 8 (6.7) | 1 (5.0) | 9 (6.5) | | |

**Supplementary Table S3.** Relationship between stunting and socio-economic factors (n=200)

| Sociodemographic factors | Normal n (%) | Stunting n (%) | Total | Test* | P value |
|---|---|---|---|---|---|
| **Was the family displaced due to war?** | | | | Fisher's exact test | 0.177 |
| Yes | 166 (94.9) | 22 (88.0) | 188 (94.0) | | |
| No | 9 (5.1) | 3 (12.0) | 12 (6.0) | | |
| **Does the family face food shortages?** | | | | Pearson's $\chi^2$ = 0.458 | 0.498 |
| Yes | 117 (66.9) | 15 (60.0) | 132 (66.0) | | |
| No | 58 (33.1) | 10 (40.0) | 68 (34.0) | | |
| **Does the family receive food aid?** | | | | Pearson's $\chi^2$ = 0.049 | 0.826 |
| Yes | 67 (38.3) | 9 (36.0) | 76 (38.0) | | |
| No | 108 (61.7) | 16 (64.0) | 124 (62.0) | | |
| **Source of drinking water** | | | | Fisher's exact test | 0.459 |
| Municipal | 77 (44.0) | 11 (44.0) | 88 (44.0) | | |
| Tanker | 60 (34.3) | 8 (32.0) | 68 (34.0) | | |
| Bottled | 30 (17.1) | 3 (12.0) | 33 (16.5) | | |
| Others | 8 (4.6) | 3 (12.0) | 11 (5.5) | | |

*Note.* P values are two-sided. Pearson's $\chi^2$ test was used unless expected cell counts were <5, in which case Fisher's exact test was used ([a]).

**Supplementary Table S4.** Relationship between stunting and the child's age

| Child age | Normal n (%) | Stunting n (%) | Total | Test | P value |
|---|---|---|---|---|---|
| 6 months | 35 (21.2) | 10 (40.0) | 45 (23.7) | Pearson's $\chi^2$ = 6.306 | 0.177 |
| 7–9 months | 33 (20.0) | 6 (24.0) | 39 (20.5) | | |
| 10–12 months | 28 (17.0) | 4 (16.0) | 32 (16.8) | | |
| 13–18 months | 38 (23.0) | 2 (8.0) | 40 (21.1) | | |

| | | | |
|---|---|---|---|
| 19–24 months | 31 (18.8) | 3 (12.0) | 34 (17.9) |

*Note. Percentages in the Normal and Stunting columns are column percentages within nutritional-status category; percentages in the Total column are based on the full age-analysis denominator (n=190), restricted to children whose recorded ages fell within the prespecified 6-24-month bands.*

**Supplementary Table S5.** Relationship between stunting and child demographic factors (n=200)

| Demographic factors | Normal n (%) | Stunting n (%) | Total | Test | P value |
|---|---|---|---|---|---|
| **Any chronic disease** | | | | **Fisher's exact test** | 1.000 |
| Yes | 8 (4.6) | 1 (4.0) | 9 (4.5) | | |
| No | 167 (95.4) | 24 (96.0) | 191 (95.5) | | |
| **Has the child been treated for malnutrition previously?** | | | | **Pearson's $\chi^2$ = 0.306** | 0.580 |
| Yes | 66 (37.7) | 8 (32.0) | 74 (37.0) | | |
| No | 109 (62.3) | 17 (68.0) | 126 (63.0) | | |

**Supplementary Table S6.** Relationship between underweight and socio-economic factors (n=199)

| Sociodemographic factors | Normal n (%) | Underweight n (%) | Total | Test | P value |
|---|---|---|---|---|---|
| **Mothers' education** | | | | Pearson's $\chi^2$ = 0.487 | 0.485 |
| Preparatory or less | 21 (13.2) | 7 (17.5) | 28 (14.1) | | |
| Secondary or more | 138 (86.8) | 33 (82.5) | 171 (85.9) | | |
| **Fathers' education** | | | | Pearson's $\chi^2$ = 0.001 | 0.981 |
| Preparatory or less | 48 (30.2) | 12 (30.0) | 60 (30.2) | | |
| Secondary or more | 111 (69.8) | 28 (70.0) | 139 (69.8) | | |
| **Number of children in the child's family** | | | | Pearson's $\chi^2$ = 0.006 | 0.938 |
| One child | 27 (17.0) | 7 (17.5) | 34 (17.1) | | |
| Two children or more | 132 (83.0) | 33 (82.5) | 165 (82.9) | | |
| **Child sex** | | | | Pearson's $\chi^2$ = 2.557 | 0.110 |
| Male | 90 (56.6) | 17 (42.5) | 107 (53.8) | | |
| Female | 69 (43.4) | 23 (57.5) | 92 (46.2) | | |
| **Father's employment status** | | | | Pearson's $\chi^2$ = 0.140 | 0.708 |
| Yes | 32 (20.1) | 7 (17.5) | 39 (19.6) | | |
| No | 127 (79.9) | 33 (82.5) | 160 (80.4) | | |
| **Mothers' employment status** | | | | Fisher's exact test | 1.000 |
| Yes | 3 (1.9) | 1 (2.5) | 4 (2.0) | | |
| No | 156 (98.1) | 39 (97.5) | 195 (98.0) | | |

| Income (n = 138) | | | | Fisher's exact test | 1.000 |
|---|---|---|---|---|---|
| less than 1000 NIS | 106 (93.0) | 23 (95.8) | 129 (93.5) | | |
| 1000 NIS or more | 8 (7.0) | 1 (4.2) | 9 (6.5) | | |

**Supplementary Table S7.** Relationship between underweight and socio-economic factors (n=199)

| Sociodemographic factors | Normal n (%) | Underweight n (%) | Total | Test | P value |
|---|---|---|---|---|---|
| **Was the family displaced due to war?** | | | | **Pearson's $\chi^2$ = 0.191** | 0.662 |
| Yes | 150 (94.3) | 37 (92.5) | 187 (94.0) | | |
| No | 9 (5.7) | 3 (7.5) | 12 (6.0) | | |
| **Does the family face food shortages?** | | | | **Pearson's $\chi^2$ = 6.185** | **0.013*** |
| Yes | 98 (61.6) | 33 (82.5) | 131 (65.8) | | |
| No | 61 (38.4) | 7 (17.5) | 68 (34.2) | | |
| **Does the family receive food aid?** | | | | **Pearson's $\chi^2$ = 0.574** | 0.449 |
| Yes | 62 (39.0) | 13 (32.5) | 75 (37.7) | | |
| No | 97 (61.0) | 27 (67.5) | 124 (62.3) | | |
| **Source of drinking water** | | | | **Pearson's $\chi^2$ = 4.712** | 0.194 |
| Municipal | 71 (44.7) | 17 (42.5) | 88 (44.2) | | |
| Tanker | 55 (34.6) | 12 (30.0) | 67 (33.7) | | |
| Bottled | 27 (17.0) | 6 (15.0) | 33 (16.6) | | |
| Others | 6 (3.8) | 5 (12.5) | 11 (5.5) | | |

**Statistically significant*

**Supplementary Table S8.** Relationship between underweight and child's age

| Child age | Normal n (%) | Underweight n (%) | Total | Test | P value |
|---|---|---|---|---|---|
| 6 months | 39 (25.7) | 6 (16.2) | 45 (23.8) | Pearson's $\chi^2$ = 7.111 | 0.130 |
| 7–9 months | 27 (17.8) | 12 (32.4) | 39 (20.6) | | |
| 10–12 months | 25 (16.4) | 7 (18.9) | 32 (16.9) | | |
| 13–18 months | 36 (23.7) | 4 (10.8) | 40 (21.2) | | |
| 19–24 months | 25 (16.4) | 8 (21.6) | 33 (17.5) | | |

*Note. Percentages in the Normal and Underweight columns are column percentages within nutritional-status category; percentages in the Total column are based on the full age-analysis denominator (n=189), restricted to children whose recorded ages fell within the prespecified 6-24-month bands and for whom an underweight classification was available.*

**Supplementary Table S9.** Relationship between underweight and the child's demographic factors

| Demographic factors | Normal n (%) | Underweight n (%) | Total | Test | P value |
|---|---|---|---|---|---|
| **Any chronic disease** | | | | Fisher's exact test | 1.000 |
| Yes | 7 (4.4) | 2 (5.0) | 9 (4.5) | | |
| No | 152 (95.6) | 38 (95.0) | 190 (95.5) | | |

| **Has the child been treated for malnutrition previously?** | | | | Pearson's $\chi^2$ = 9.340 | **0.002*** |
|---|---|---|---|---|---|
| Yes | 50 (31.4) | 23 (57.5) | 73 (36.7) | | |
| No | 109 (68.6) | 17 (42.5) | 126 (63.3) | | |

**Statistically significant*

**Supplementary Table S10.** Relationship between wasting and socio-economic factors

| Sociodemographic factor | Normal n (%) | Wasting n (%) | Total | Preferred test | P value |
|---|---|---|---|---|---|
| **Mothers' education** | | | | Fisher's exact test | 0.340 |
| Preparatory or less | 5 (11.9) | 3 (27.3) | 8 (15.1) | | |
| Secondary or more | 37 (88.1) | 8 (72.7) | 45 (84.9) | | |
| **Fathers' education** | | | | Fisher's exact test | 0.711 |
| Preparatory or less | 11 (26.2) | 2 (18.2) | 13 (24.5) | | |
| Secondary or more | 31 (73.8) | 9 (81.8) | 40 (75.5) | | |
| **Number of children in the child's family** | | | | Fisher's exact test | 0.667 |
| One child | 9 (21.4) | 1 (9.1) | 10 (18.9) | | |
| Two children or more | 33 (78.6) | 10 (90.9) | 43 (81.1) | | |
| **Child sex** | | | | Fisher's exact test | 0.175 |
| Male | 19 (45.2) | 8 (72.7) | 27 (50.9) | | |
| Female | 23 (54.8) | 3 (27.3) | 26 (49.1) | | |
| **Father's employment status** | | | | Fisher's exact test | 1.000 |
| Yes | 10 (23.8) | 3 (27.3) | 13 (24.5) | | |
| No | 32 (76.2) | 8 (72.7) | 40 (75.5) | | |
| **Mothers' employment status** | | | | Fisher's exact test | 0.375 |
| Yes | 1 (2.4) | 1 (9.1) | 2 (3.8) | | |
| No | 41 (97.6) | 10 (90.9) | 51 (96.2) | | |
| **Income (n = 39)** | | | | Fisher's exact test | 0.141 |
| Less than 1000 NIS | 30 (93.8) | 5 (71.4) | 35 (89.7) | | |
| 1000 NIS or more | 2 (6.2) | 2 (28.6) | 4 (10.3) | | |

*Note. Wasting analyses were based on children with an available wasting classification. Income was available for 39 children in this subset.*

**Supplementary Table S11.** Relationship between wasting and socio-economic factors (n = 53 with available wasting classification)

| Sociodemographic factors | Normal n (%) | Wasting n (%) | Total | Test | P value |
|---|---|---|---|---|---|
| **Was the family displaced due to war?** | | | | Fisher's exact test | 1.000 |
| Yes | 39 (92.9) | 11 (100.0) | 50 (94.3) | | |
| No | 3 (7.1) | 0 (0.0) | 3 (5.7) | | |
| **Does the family face food shortages?** | | | | Fisher's exact test | 0.313 |
| Yes | 18 (42.9) | 7 (63.6) | 25 (47.2) | | |
| No | 24 (57.1) | 4 (36.4) | 28 (52.8) | | |
| **Does the family receive food aid?** | | | | Fisher's exact test | 1.000 |
| Yes | 14 (33.3) | 3 (27.3) | 17 (32.1) | | |
| No | 28 (66.7) | 8 (72.7) | 36 (67.9) | | |
| **Source of drinking water** | | | | Pearson's $\chi^2$ = **1.168** | 0.761 |
| Municipal | 19 (45.2) | 4 (36.4) | 23 (43.4) | | |
| Tanker | 14 (33.3) | 3 (27.3) | 17 (32.1) | | |
| Bottled | 6 (14.3) | 3 (27.3) | 9 (17.0) | | |
| Others | 3 (7.1) | 1 (9.1) | 4 (7.5) | | |

**Supplementary Table S12.** Relationship between wasting and the child's age (n=51)

| Child age | Normal n (%) | Wasting n (%) | Total | Test | P value |
|---|---|---|---|---|---|
| 6 months | 15 (36.6) | 1 (10.0) | 16 (31.4) | Fisher's exact test | **0.0024*** |
| 7–9 months | 13 (31.7) | 0 (0.0) | 13 (25.5) | | |
| 10–12 months | 7 (17.1) | 2 (20.0) | 9 (17.6) | | |
| 13–18 months | 4 (9.8) | 5 (50.0) | 9 (17.6) | | |
| 19–24 months | 2 (4.9) | 2 (20.0) | 4 (7.8) | | |

**Statistically significant. Note. Percentages in the Normal and Wasting columns are column percentages within nutritional-status category; percentages in the Total column are based on the full age-analysis denominator (n=51). Age-category analysis was restricted to children with an available wasting classification and a recorded age falling within the prespecified 6-24-month bands. Fisher's exact test was used.*

**Supplementary Table S13**. Relationship between wasting and child demographic factors (n=53 with available wasting classification)

| Demographic factors | Normal n (%) | Wasting n (%) | Total | Test | P value |
|---|---|---|---|---|---|
| **Any chronic disease** | | | | | |
| Yes | 2 (4.8) | 1 (9.1) | 3 (5.7) | Fisher's exact test | 0.510 |
| No | 40 (95.2) | 10 (90.9) | 50 (94.3) | | |
| **Has the child been treated for malnutrition previously?** | | | | | |
| Yes | 12 (28.6) | 3 (27.3) | 15 (28.3) | Fisher's exact test | 1.000 |
| No | 30 (71.4) | 8 (72.7) | 38 (71.7) | | |

**Supplementary Table S14.** Relationship between anaemia and socio-economic factors (n=55)

| Sociodemographic factors | Anaemia n (%) | Normal n (%) | Total | Test | P value |
|---|---|---|---|---|---|
| **Mothers' education** | | | | Fisher's exact test | 0.443 |
| Preparatory or less | 4 (11.4) | 4 (20.0) | 8 (14.5) | | |
| Secondary or more | 31 (88.6) | 16 (80.0) | 47 (85.5) | | |
| **Fathers' education** | | | | Fisher's exact test | 0.359 |
| Preparatory or less | 12 (34.3) | 4 (20.0) | 16 (29.1) | | |
| Secondary or more | 23 (65.7) | 16 (80.0) | 39 (70.9) | | |
| **Number of children in the child's family** | | | | Fisher's exact test | 0.469 |
| One child | 5 (14.3) | 5 (25.0) | 10 (18.2) | | |
| Two children or more | 30 (85.7) | 15 (75.0) | 45 (81.8) | | |
| **Child sex** | | | | Fisher's exact test | 0.091 |
| Male | 23 (65.7) | 8 (40.0) | 31 (56.4) | | |
| Female | 12 (34.3) | 12 (60.0) | 24 (43.6) | | |
| **Father's employment status** | | | | Fisher's exact test | **0.020*** |
| Yes | 4 (11.4) | 8 (40.0) | 12 (21.8) | | |
| No | 31 (88.6) | 12 (60.0) | 43 (78.2) | | |
| **Mothers' employment status** | | | | Fisher's exact test | 1.000 |
| Yes | 1 (2.9) | 1 (5.0) | 2 (3.6) | | |
| No | 34 (97.1) | 19 (95.0) | 53 (96.4) | | |
| **Income (n=47)** | | | | Fisher's exact test | 0.126 |
| Less than 1000 NIS | 30 (100.0) | 15 (88.2) | 45 (95.7) | | |
| 1000 NIS or more | 0 (0.0) | 2 (11.8) | 2 (4.3) | | |

**Statistically significant. Note. Anaemia was defined from the numeric haemoglobin value as Hb <11 g/dL among the 55 haemoglobin-tested children.*

**Supplementary Table S15.** Relationship between anaemia and socio-economic factors (n=55)

| Sociodemographic factors | Anaemia n (%) | Normal n (%) | Total | Test | P value |
|---|---|---|---|---|---|
| **Was the family displaced due to war?** | | | | Fisher's exact test | 0.131 |

| | | | | | |
|---|---|---|---|---|---|
| Yes | 34 (97.1) | 17 (85.0) | 51 (92.7) | | |
| No | 1 (2.9) | 3 (15.0) | 4 (7.3) | | |
| **Does the family face food shortages?** | | | | Fisher's exact test | 1.000 |
| Yes | 27 (77.1) | 15 (75.0) | 42 (76.4) | | |
| No | 8 (22.9) | 5 (25.0) | 13 (23.6) | | |
| **Does the family receive food aid?** | | | | Fisher's exact test | 0.572 |
| Yes | 16 (45.7) | 7 (35.0) | 23 (41.8) | | |
| No | 19 (54.3) | 13 (65.0) | 32 (58.2) | | |
| **Source of drinking water** | | | | Pearson's $\chi^2$ = 2.611 | 0.456 |
| Municipal | 11 (31.4) | 7 (35.0) | 18 (32.7) | | |
| Tanker | 15 (42.9) | 5 (25.0) | 20 (36.4) | | |
| Bottled | 8 (22.9) | 6 (30.0) | 14 (25.5) | | |
| Others | 1 (2.9) | 2 (10.0) | 3 (5.5) | | |

*Note. Anaemia was defined from the numeric haemoglobin value as Hb <11 g/dL among the 55 haemoglobin-tested children.*

**Supplementary Table S16.** Relationship between anaemia and child's age (n=55)

| Child age | Anaemia n (%) | Normal n (%) | Total | Test | P value |
|---|---|---|---|---|---|
| 6 months | 5 (14.3) | 0 (0.0) | 5 (9.1) | Fisher's exact test | 0.092 |
| 7-9 months | 3 (8.6) | 0 (0.0) | 3 (5.5) | | |
| 10-12 months | 2 (5.7) | 3 (15.0) | 5 (9.1) | | |
| 13-18 months | 14 (40.0) | 13 (65.0) | 27 (49.1) | | |
| 19-24 months | 11 (31.4) | 4 (20.0) | 15 (27.3) | | |

*Note. Percentages in the Anaemia and Normal columns are column percentages within anaemia-status category; percentages in the Total column are based on the full age-analysis denominator (n=55). Anaemia was defined from the numeric haemoglobin value as Hb <11 g/dL among the 55 haemoglobin-tested children.*

**Supplementary Table S17.** Relationship between anaemia and children's demographic factors (n=55)

| Demographic factors | Anaemia n (%) | Normal n (%) | Total | Test | P value |
|---|---|---|---|---|---|
| **Any chronic disease** | | | | Fisher's exact test | 1.000 |
| Yes | 2 (5.7) | 1 (5.0) | 3 (5.5) | | |
| No | 33 (94.3) | 19 (95.0) | 52 (94.5) | | |
| **Has the child been treated for malnutrition previously?** | | | | Fisher's exact test | 0.158 |
| Yes | 16 (45.7) | 5 (25.0) | 21 (38.2) | | |
| No | 19 (54.3) | 15 (75.0) | 34 (61.8) | | |

*Note. Anaemia was defined from the numeric haemoglobin value as Hb <11 g/dL among the 55 haemoglobin-tested children.*

**Supplementary Table S18.** Children's dietary intake and feeding practices of children attending the PHCC in the Gaza Strip (n=200).

| Feeding practices | n | % |
|---|---|---|

| | | |
|---|---|---|
| **Was the child breastfed?** | | |
| Yes | 172 | 86.0 |
| No | 28 | 14.0 |
| **Feeding methods** | | |
| Breastfeeding only | 76 | 38.0 |
| Bottle only | 19 | 9.5 |
| Mixed | 105 | 52.5 |
| **Has complementary feeding started?** | | |
| Yes | 170 | 85.0 |
| No | 30 | 15.0 |
| **Age started (months), Mean ± SD (Min-Max), n=166** | **5.9±2.1 (1–22)** | |
| Less than 4 months | 3 | 1.8 |
| 4–5 months | 53 | 31.9 |
| 6 months | 85 | 51.2 |
| 7–8 months | 15 | 9.0 |
| 9 months and more | 10 | 6.0 |
| **First type of food introduced (Multiple response question)** | | |
| Juice | 27 | 13.5 |
| Cereals | 75 | 37.5 |
| Vegetables | 76 | 38.0 |
| Fruits | 60 | 30.0 |
| Others | 35 | 17.5 |
| **Has breastfeeding stopped?** | | |
| Yes | 59 | 29.5 |
| No | 141 | 70.5 |
| **Age of stopping breastfeeding by months, Mean ± SD (Min-Max)** | **12.0±5.7 (1–24)** | |
| **Number of meals per day, Mean ± SD (Min-Max)** | **2.9±1.4 (1–12)** | |
| **Food groups consumed in the last 24 hours (Multiple response question)** | | |
| Cereals | 54 | 27.0 |
| Vegetables | 81 | 40.5 |
| Fruit | 68 | 34.0 |
| Meat & legumes | 14 | 7.0 |
| Dairy | 108 | 54.0 |
| Sweets | 17 | 8.5 |
| **Do you add micronutrient supplements?** | | |
| Yes | 107 | 53.5 |
| No | 93 | 46.5 |
| **Feeding difficulties in the last week?** | | |
| Yes | 94 | 47.0 |
| No | 106 | 53.0 |

*Note. One child had a recorded complementary-feeding start age of 6.3 months and was grouped within the 6-month category for presentation.*

**Supplementary Table S19.** Role of PHCC nurses in addressing child malnutrition through mothers' instructions as reported by mothers of children (n=200)

| Variables | n | % |
|---|---|---|
| **Do you visit the child health center regularly?** | | |
| Yes | 156 | 78.0 |
| No | 44 | 22.0 |
| **Do nurses at PHCC provide nutrition advice?** | | |
| Yes | 142 | 71.0 |

| | | |
|---|---|---|
| No | 58 | 29.0 |
| **Type of guidance (Breastfeeding)** | | |
| Yes | 101 | 50.5 |
| No | 99 | 49.5 |
| **Type of guidance (Complementary feeding)** | | |
| Yes | 56 | 28.0 |
| No | 144 | 72.0 |
| **Type of guidance (Food hygiene)** | | |
| Yes | 32 | 16.0 |
| No | 168 | 84.0 |
| **Type of guidance (information about anaemia and supplements)** | | |
| Yes | 23 | 11.5 |
| No | 177 | 88.5 |
| **Type of guidance (No guidance provided at all)** | | |
| Yes | 42 | 21.0 |
| No | 158 | 79.0 |
| **Do you believe nurses play a role in improving your child's nutrition?** | | |
| Yes | 157 | 78.5 |
| No | 43 | 21.5 |
| **Did the nurse assess your child's growth charts with you?** | | |
| Yes | 105 | 52.5 |
| No | 95 | 47.5 |
| **Did you receive written or visual educational materials?** | | |
| Yes | 68 | 34.0 |
| No | 132 | 66.0 |

**Supplementary Table S20.** Women's level of knowledge on Breastfeeding & Complementary /Feeding

| Statements | N | % |
|---|---|---|
| **Colostrum is beneficial and should be given to the infant.** | | |
| Yes | 169 | 84.5 |
| No | 31 | 15.5 |
| **Exclusive breastfeeding means giving only breast milk without water or herbs until 6 months.** | | |
| Yes | 135 | 67.5 |
| No | 65 | 32.5 |
| **Good signs of milk adequacy include sufficient urination and weight gain.** | | |
| Yes | 192 | 96.0 |
| No | 8 | 4.0 |
| **It is preferable to reduce breastfeeding when the infant is ill.*** | | |
| Yes | 55 | 27.5 |
| No | 145 | 72.5 |
| **Correct positioning and latch reduce nipple cracks and engorgement.** | | |
| Yes | 176 | 88.0 |
| No | 24 | 12.0 |
| **Breastfeeding alongside food is recommended until 24 months or more.** | | |
| Yes | 171 | 85.5 |
| No | 29 | 14.5 |
| **Complementary feeding should start at 6 months with continued breastfeeding.** | | |
| Yes | 192 | 96.0 |
| No | 8 | 4.0 |
| **Food consistency should progress from smooth purees to more solid textures as the child ages.** | | |
| Yes | 190 | 95.0 |

| | | |
|---|---|---|
| No | 10 | 5.0 |
| **Sugar/salt should be avoided in infant food.** | | |
| Yes | 187 | 93.5 |
| No | 13 | 6.5 |
| **Food hygiene and safety are not necessary at this age.*** | | |
| Yes | 30 | 15.0 |
| No | 170 | 85.0 |
| **Sugary drinks and energy drinks are suitable for infants.*** | | |
| Yes | 15 | 7.5 |
| No | 185 | 92.5 |

*Note.* Correct answers are highlighted. *Reverse coded items.

**Supplementary Table S21.** Women's level of practice on Breastfeeding & Complementary /Feeding

| Practice statements | Never | Rarely | Sometimes | Often | Always | Mean | SD | Rank |
|---|---|---|---|---|---|---|---|---|
| I started breastfeeding my baby within the first hour after birth. | 12 (6) | 8 (4) | 2 (1) | 15 (7.5) | 163 (81.5) | 4.5 | 1.1 | 3 |
| I practised exclusive breastfeeding until 6 months without water/herbs/formula. | 21 (10.5) | 11 (5.5) | 30 (15) | 33 (16.5) | 105 (52.5) | 4.0 | 1.4 | 11 |
| I used a pacifier with my baby during the first six months.* | 125 (62.5) | 12 (6) | 14 (7) | 20 (10) | 29 (14.5) | 4.2 | 1.4 | 8 |
| I increase or maintain breastfeeding frequency when my baby is ill. | 7 (3.5) | 16 (8) | 20 (10) | 27 (13.5) | 130 (65) | 4.3 | 1.1 | 6 |
| I check my baby's positioning and latch during breastfeeding. | 7 (3.5) | 6 (3) | 9 (4.5) | 19 (9.5) | 159 (79.5) | 4.6 | 1.0 | 2 |
| I continued breastfeeding alongside complementary feeding after 12 months. | 23 (11.5) | 5 (2.5) | 16 (8) | 26 (13) | 130 (65) | 4.2 | 1.4 | 9 |
| I started complementary feeding for my baby at 6 months. | 10 (5) | 13 (6.5) | 11 (5.5) | 29 (14.5) | 137 (68.5) | 4.3 | 1.2 | 5 |
| I gradually increase my child's food consistency as they get older. | 7 (3.5) | 14 (7) | 13 (6.5) | 17 (8.5) | 149 (74.5) | 4.4 | 1.1 | 4 |
| I regularly provide iron-rich or fortified foods. | 10 (5) | 16 (8) | 20 (10) | 21 (10.5) | 133 (66.5) | 4.3 | 1.2 | 7 |
| I avoid adding sugar/salt to my child's food or keep them to a minimum. | 18 (9) | 18 (9) | 29 (14.5) | 20 (10) | 115 (57.5) | 4.0 | 1.4 | 10 |
| I wash my hands with safe water and use clean utensils when preparing/feeding the child. | 2 (1) | 6 (3) | 5 (2.5) | 11 (5.5) | 176 (88) | 4.8 | 0.7 | 1 |
| I avoid giving sugary/soft/energy drinks to my child. | 31 (15.5) | 15 (7.5) | 17 (8.5) | 15 (7.5) | 122 (61) | 3.9 | 1.5 | 12 |

*Reverse coded item.

**Supplementary Table S22.** Descriptive analysis of Knowledge and practice scores

| Domain | Mean ± SD | Min-Max | Lowest-Highest possible scores |
|---|---|---|---|
| **Knowledge** | 9.5 ± 1.4 | 5-11 | 0-11 |
| **Practice** | 51.5 ± 7.5 | 20-60 | 12-60 |

**Note: Knowledge score based on 11 retained items after reliability testing, possible range 0–11. Practice score based on 12 retained items after reliability testing, possible range 12–60.*

**Supplementary Table S23.** Correlation between women's knowledge and practice scores on breastfeeding and complementary feeding

| Variable | Practice score | Knowledge score |
|---|---|---|

| | | |
|---|---|---|
| **Practice score** | 1 | 0.177* |
| **Sig. (2-tailed)** | | 0.012 |
| **N** | 200 | 200 |
| **Knowledge score** | 0.177* | 1 |
| **Sig. (2-tailed)** | 0.012 | |
| **N** | 200 | 200 |

**Note. Statistically significant at 0.05.*

**Supplementary Table S24**. The relationship between women's level of knowledge and the children's nutritional status

| Nutritional status | Number | Knowledge mean ± SD | t | P value |
|---|---|---|---|---|
| **Stunting** | | | | |
| Stunting | 25 | 9.8 ± 1.2 | 0.990 | 0.324 |
| Normal | 175 | 9.5 ± 1.5 | | |
| **Underweight** | | | | |
| Underweight | 40 | 9.5 ± 1.5 | -0.161 | 0.873 |
| Normal | 159 | 9.5 ± 1.4 | | |
| **Wasting** | | | | |
| Wasting | 11 | 10.55 ± 0.69 | 1.553 | 0.127 |
| Normal | 42 | 9.98 ± 1.16 | | |
| **Anaemia (n=55)** | | | | |
| 11 or more (not anaemic) | 20 | 9.85 ± 1.04 | -0.451 | 0.654 |
| less than 11 (anaemic) | 35 | 10.00 ± 1.26 | | |

*Note. Anaemia was defined from the numeric haemoglobin value as Hb <11 g/dL among the 55 haemoglobin-tested children.*

**Supplementary Table S25**. The relationship between women's level of practice and the children's nutritional status

| Nutritional status | Number | Practice mean ± SD | t | P value |
|---|---|---|---|---|
| **Stunting** | | | | |
| Stunting | 25 | 49.3 ± 10.4 | -1.543 | 0.124 |
| Normal | 175 | 51.8 ± 7.0 | | |
| **Underweight** | | | | |
| Underweight | 40 | 51.2 ± 6.4 | -0.315 | 0.753 |
| Normal | 159 | 51.6 ± 7.7 | | |
| **Wasting** | | | | |
| Wasting | 11 | 52.18 ± 5.78 | 0.637 | 0.527 |
| Normal | 42 | 50.36 ± 8.99 | | |
| **Anaemia (n=55)** | | | | |
| Not anaemic | 20 | 51.30 ± 6.91 | -0.384 | 0.702 |
| Anaemic | 35 | 51.97 ± 5.82 | | |

*Note. Anaemia was defined from the numeric haemoglobin value as Hb <11 g/dL among the 55 haemoglobin-tested children.*

**Supplementary Table S26.** Full adjusted logistic regression models for stunting and underweight

| Predictor | Stunting knowledge model aOR (95% CI) | p-value | Stunting practice model aOR (95% CI) | p-value | Underweight knowledge model aOR (95% CI) | p-value | Underweight practice model aOR (95% CI) | p-value |
|---|---|---|---|---|---|---|---|---|
| Younger age (6-12 vs 13-24 months) | **2.93 (1.01 to 8.49)** | **0.048** | 2.88 (1.00 to 8.30) | 0.050 | 2.02 (0.88 to 4.62) | 0.098 | 1.95 (0.86 to 4.43) | 0.109 |
| Male sex | 0.90 (0.38 to 2.12) | 0.802 | 0.99 (0.42 to 2.37) | 0.989 | 0.60 (0.28 to 1.27) | 0.184 | 0.61 (0.29 to 1.30) | 0.203 |

| | | | | | | | | |
|---|---|---|---|---|---|---|---|---|
| Previous malnutrition treatment | 1.00 (0.39 to 2.58) | 0.995 | 0.95 (0.37 to 2.46) | 0.923 | **3.56 (1.61 to 7.87)** | **0.002** | **3.49 (1.59 to 7.66)** | **0.002** |
| Household adversity index (per 1-point increase) | 0.93 (0.60 to 1.44) | 0.737 | 0.86 (0.55 to 1.35) | 0.520 | 1.10 (0.75 to 1.63) | 0.626 | 1.08 (0.73 to 1.61) | 0.689 |
| Maternal knowledge score (per 1-point increase) | 1.20 (0.86 to 1.66) | 0.284 | | | 1.09 (0.83 to 1.43) | 0.537 | | |
| Maternal practice score (per 1-point increase) | | | 0.96 (0.91 to 1.01) | 0.103 | | | 0.99 (0.94 to 1.04) | 0.620 |

**Statistically significant at 0.05. Note. The household adversity index assigned one point each for war-related displacement, household food shortage, receipt of food aid, non-municipal drinking water, and paternal unemployment. Given the limited number of outcome events, these adjusted models should be interpreted as exploratory. Stunting models were based on 190 children whose recorded ages fell within the prespecified 6-24-month bands. Underweight models were based on 189 children whose recorded ages fell within these bands and for whom an underweight classification was available.*

**Supplementary Table S27**. Secondary linear regression analyses of maternal knowledge and practice scores

| Predictor | Knowledge score β (95% CI) | p-value | Practice score β (95% CI) | p-value |
|---|---|---|---|---|
| Nurse nutrition advice | 0.33 (-0.13 to 0.79) | 0.162 | 1.81 (-0.60 to 4.22) | 0.139 |
| Growth-chart review | -0.23 (-0.64 to 0.18) | 0.274 | 0.70 (-1.46 to 2.87) | 0.524 |
| Written/visual educational materials | **0.45 (0.02 to 0.89)** | **0.040** | 0.14 (-2.14 to 2.43) | 0.902 |
| Regular PHCC attendance | -0.05 (-0.54 to 0.44) | 0.841 | -0.04 (-2.60 to 2.52) | 0.974 |
| Maternal knowledge score (per 1-point increase) | | | **0.86 (0.13 to 1.60)** | **0.022** |

**Statistically significant at 0.05. Note. The knowledge model examined associations of PHCC service variables with maternal knowledge score. The practice model examined associations of the same PHCC variables and maternal knowledge score with maternal practice score.*

**Supplementary Table S28.** Sensitivity analyses replacing the adversity index with household food shortage

| Predictor | Stunting knowledge model aOR (95% CI) | p-value | Stunting practice model aOR (95% CI) | p-value | Underweight knowledge model aOR (95% CI) | p-value | Underweight practice model aOR (95% CI) | p-value |
|---|---|---|---|---|---|---|---|---|
| Younger age (6-12 vs 13-24 months) | **2.96 (1.02 to 8.58)** | **0.046*** | 2.88 (1.00 to 8.33) | 0.050 | 2.22 (0.96 to 5.16) | 0.063 | 2.11 (0.92 to 4.83) | 0.076 |
| Male sex | 0.89 (0.38 to 2.09) | 0.787 | 0.98 (0.41 to 2.34) | 0.965 | 0.60 (0.28 to 1.28) | 0.189 | 0.61 (0.29 to 1.31) | 0.206 |
| Previous malnutrition treatment | 0.99 (0.38 to 2.57) | 0.980 | 0.96 (0.37 to 2.49) | 0.926 | **3.20 (1.45 to 7.10)** | **0.004*** | **3.14 (1.43 to 6.92)** | **0.004** |
| Household food shortage | 0.91 (0.36 to 2.29) | 0.844 | 0.78 (0.31 to 1.98) | 0.604 | 2.30 (0.89 to 5.93) | 0.086 | 2.16 (0.84 to 5.53) | 0.110 |
| Maternal knowledge score (per 1-point increase) | 1.19 (0.86 to 1.66) | 0.289 | | | 1.12 (0.85 to 1.47) | 0.427 | | |
| Maternal practice score (per 1-point increase) | | | 0.96 (0.91 to 1.01) | 0.110 | | | 0.99 (0.94 to 1.04) | 0.731 |

**Statistically significant at 0.05.*
